# Covid-19 predictions using a Gauss model, based on data from April 2

**DOI:** 10.1101/2020.04.06.20055830

**Authors:** Janik Schüttler, Reinhard Schlickeiser, Frank Schlickeiser, Martin Kröger

**Author notes:** Electronic address (Reinhard Schlickeiser). Electronic address (Martin Kröger). We submitted the first version to medRxiv.org (MEDRXIV/2020/055830) on April 5, 2020. This document is the second, revised version, but plots and data analysis, based on data from April 2, remain the same. We added acknowledgements.

## Abstract

We propose a Gauss model (GM), a map from time to the bell-shaped Gauss function to model the deaths per day and country, as a quick and simple model to make predictions on the coronavirus epidemic. Justified by the sigmoidal nature of a pandemic, i.e. initial exponential spread to eventual saturation, we apply the GM to existing data, as of April 2, 2020, from 25 countries during first corona pandemic wave and study the model’s predictions. We find that logarithmic daily fatalities caused by Covid-19 are well described by a quadratic function in time. By fitting the data to second order polynomials from a statistical *χ*^2^-fit with 95% confidence, we are able to obtain the characteristic parameters of the GM, i.e. a width, peak height and time of peak, for each country separately, with which we extrapolate to future times to make predictions. We provide evidence that this supposedly oversimplifying model might still have predictive power and use it to forecast the further course of the fatalities caused by Covid-19 per country, including peak number of deaths per day, date of peak, and duration within most deaths occur. While our main goal is to present the general idea of the simple modeling process using GMs, we also describe possible estimates for the number of required respiratory machines and the duration left until the number of infected will be significantly reduced.

## I. Introduction

Nowadays, numerous models to predict the spreading of infectious diseases like Covid-19 are available, for example the actively discussed susceptible-infected-removed (SIR) model [9–11]. Many of these models are either toy models that cannot make reliable predictions or they are so complex, by taking into account a wide range of factors, that simple predictions are not possible. In times of the coronavirus epidemic, predictions such as the maximum number of fatalities per day or the date of the peak number of newly seriously sick persons per day (SSPs) are valuable data for governments around the world, especially those facing the beginning of an exponential increase of casualties, and we hope to serve the people in charge with the here presented approach. In particular, *fast* predictions on the course of the coronavirus disease are crucial for policy makers to optimize their managing of the disease wave. To feed into the current debate on infectious disease models, we would like to propose a Gauss model (GM) as a simple, but effective description of fatalities caused by Covid-19 over time, similar to recent studies for the US [12] and for Germany [13]. In contrast to this previous work, we choose to use the logarithm of the reported *daily death* rates [2], instead of *cumulative infections*, as monitored input data and we also do not rely on doubling times.

The Gaussian model maps time to the bell-shaped Gauss function to fit existing data of deaths per day and country, and to use this fit to extrapolate the deaths per day to future times. Though the GM may appear too simple to be predictive, and it most likely is, we can justify its use by several arguments: 1) the GM captures the data available today well, including the entire first epidemic wave in China, and 2) epidemics are initially exponential and eventually saturating processes in cumulative quantities and thus give rise to bell-shaped daily quantities, like the Gaussian function – all of which we will explain in detail.

Please keep in mind that we are no epidemiologists and have no prior expertise in the modelling of diseases. Also, with the here presented GM we dare, by no means, to present a model capable of similar mechanistic and causal richness compared with existing infectious disease models. Our only addition is to note and use for predictions the macroscopic Gaussian nature of the time evolution of cumulative fatalities that is universal among all countries.

## II. Results

### A. Gauss model (GM)

We model the time-dependent daily change of infections and daily change of deaths with their own, a priori independent, time-dependent Gaussian functions denoted by *i*(*t*) and *d*(*t*). Each Gaussian is a bell-shaped curve, the black line in Fig. 1(a), characterized by three independent parameters: a width, a maximum height and a time at which the Gaussian curve attains this maximum height. For any value of these parameters, the *general* form of the Gauss function – the bell-shaped curve in Fig. 1(a) – is preserved, but the *concrete* fit to given data can be optimized, as illustrated in Fig. 1(b) for varying parameters.

**Fig 1:**
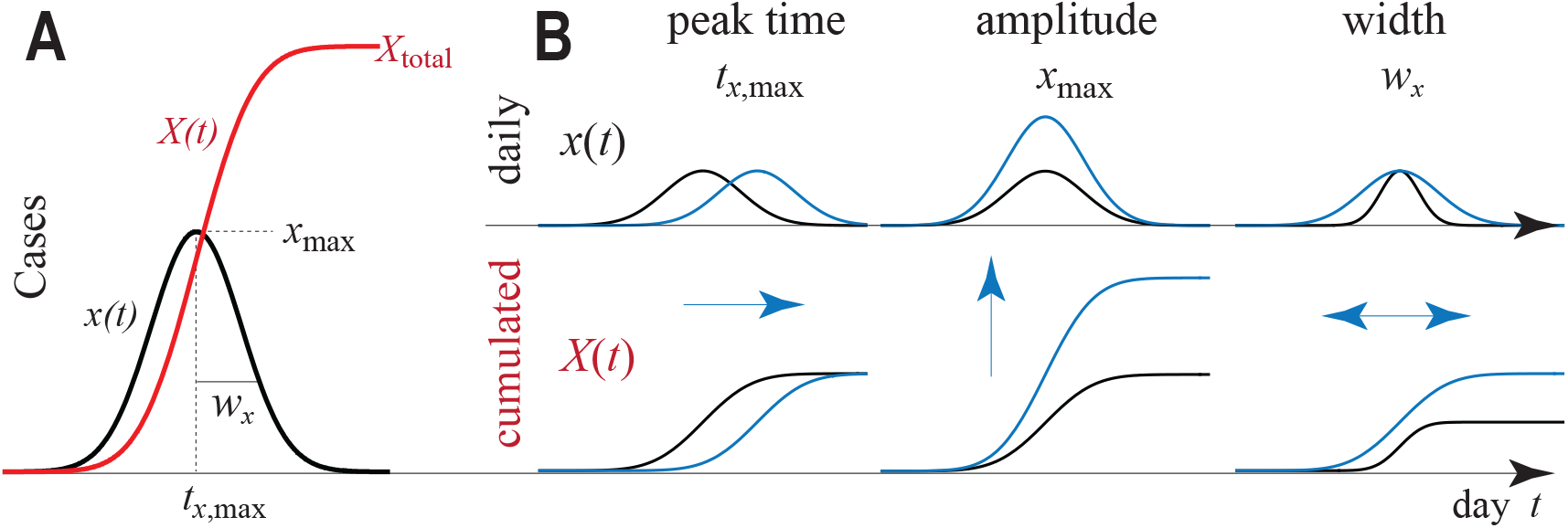
**(a)** GM for time evolution of a daily quantity *x*(*t*) (black) and the corresponding total quantity *X*(*t*) (red), which is the cumulative sum of *x*(*t*) until time *t*. **(b)** Consequences of varying the three parameters describing the GM: width *wx*, maximum height *x*max and time of maximum height *t*max for both the daily (top) and total (bottom) rates. In this work *x* stands for either deaths (*x* = *d*) or confirmed infections (*x* = *i*).

It must be emphasized that we model the *daily* change of deaths, in contrast to the *cumulative* number of deaths, more frequently available in public, because the change of deaths allow for a more stable fit around its maximum, i.e. the time of interest for predicted quantities. We will explain this point in the discussion. The cumulative deaths are the sum of all previous daily deaths up to today, while the number of daily deaths in turn is the difference of two consecutive days in cumulative deaths. In Fig. 1(a), the red plot illustrates the cumulative number of deaths as a function of time for the respective daily number of deaths in the same panel.

### B. Logarithmic daily fatalities are quadratic

Next, we fit a polynomial of second order to the logarithm of *d* of 14 countries as a function of time using a *χ*^2^-fit. The resulting quadratic fit is plotted in Fig. 2. For the remaining 11 countries, similar fits could only be performed for the daily number of infections.

**Fig 2:**
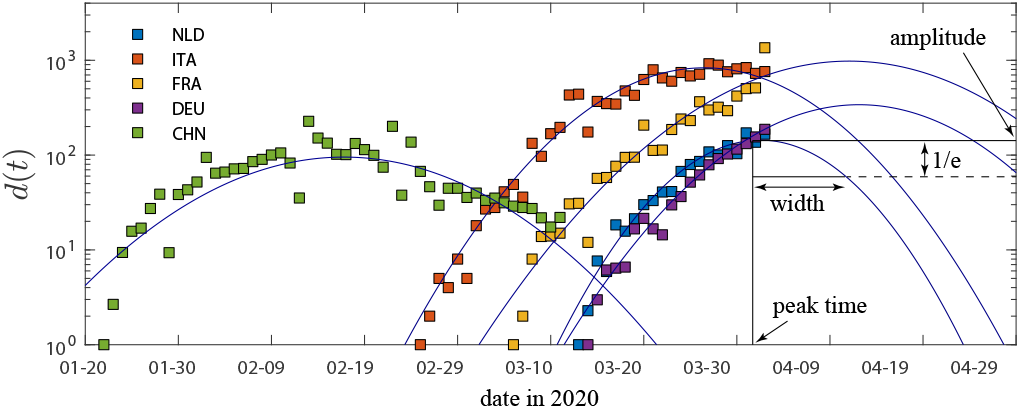
Logarithmic reported number of daily fatalities (squares) and the quadratic fits of number of daily fatalities (lines) over time for some countries. The plots demonstrate the quadratic nature of the logarithmic fatalities per day.The meaning of the GM parameters is highlighted in the inset. Raw data taken from Ref. [2].

We prefer to base any quantitative conclusions only on the number of deaths, and not on the number of infections per day. Deaths are better documented than monitored infections in nearly all countries. A death caused by Covid-19 is easier to count than an infection, which might as well cause none to moderate symptoms and hence might remain uncounted. Statistically, a constant fraction of infected die from Covid-19 at a later time after being registered as infected [4, 19]. Thus, infection and death curves are equivalent descriptions of time evolution of Covid-19, and the coefficients characterizing their shape can be expected to be closely related. To demonstrate that both, infections and deaths, follow the GM, we analyzed and show results for both measures.

### C. The fitted parameters

Using the fitted polynomial coefficients we compute the three parameters of the GM, i.e. maximum height, time of maximum height and curve width, for each country. For mathematical details, please refer to the appendix. To demonstrate the universal Gaussian nature of the daily fatalities over time *d*, we display them in Fig. 3(a,b), normalized so that all curves have unit width, maximum and time of maximum. The same plots for the cumulative fatalities *D* are shown in Fig. 3(c,d). Daily infections *i*, daily fatalities *d*, cumulative infections *I* and cumulative fatalities *D*, all fit neatly onto the unit GM curve or its cumulative function, plotted in gray in the back for reference. China, which is the only country to provide data from its first pandemic wave for times greater than 0.6 (in normalized units), fits to the GM well over the entire significant course of infections and fatalities. This sparks the hope that the used GM will have predictive power for the remaining countries also after the maximum. The fits already provide sufficient evidence that the part prior to the maximum is captured well by the GM. The resulting GM parameters are listed and plotted in Fig. 4. For most countries the GM width is within 10 and 15 days, roughly half of all countries have passed their peak of daily fatalities already and the peak is roughly below 20 fatalities per day and per million people.

**Fig 3:**
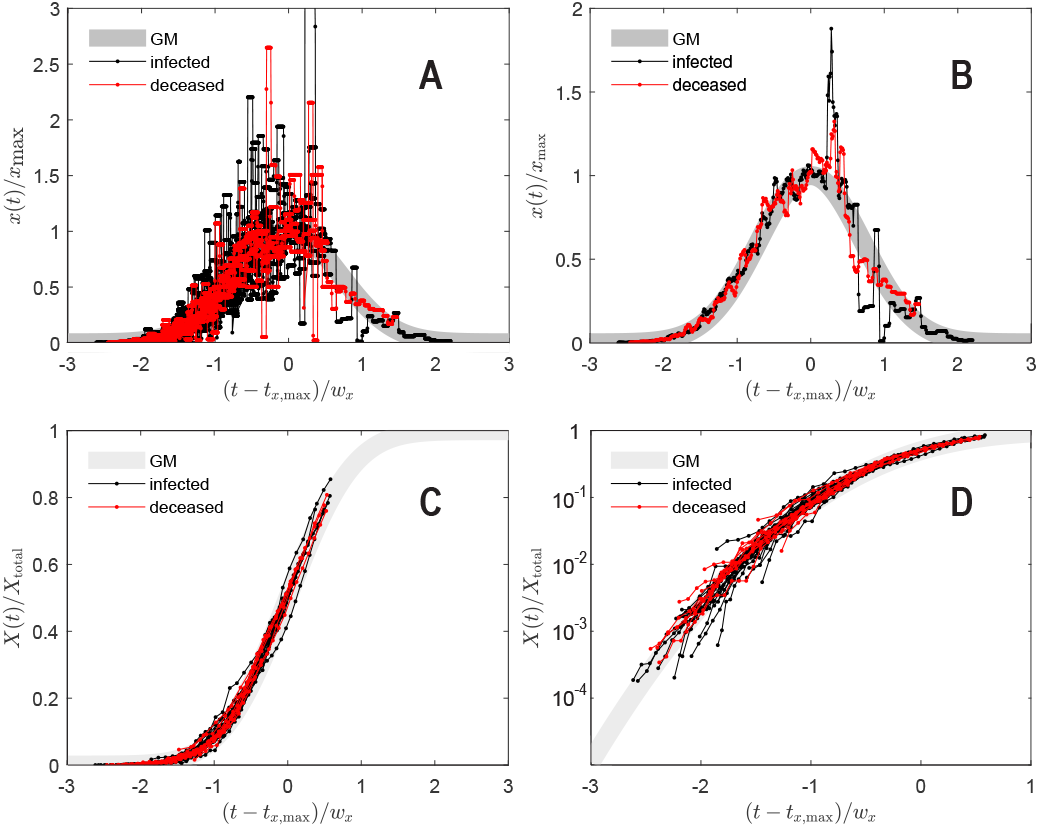
Shifted and rescaled data separately for each of the 25 countries. **(a)** number of daily infections and fatalities. **(b)** obtained from (a) by averaging over entries at same (*t − t*_*d*,max_)*/w*_*d*_ (bin size 0.01). **(c)** Number of cumulative infections and fatalities over time in normal scale, and **(d)** in logarithmic scale to appreciate different regimes with better resolution. **(a)-(d)** Thick gray is the theory expression (1) for daily, and (A7) for cumulative casualties. Data beyond (*t − t*_*d*,max_)*/w*_*d*_ = 0.6 is from China alone. Raw data taken from Ref. [2].

**Fig 4:**
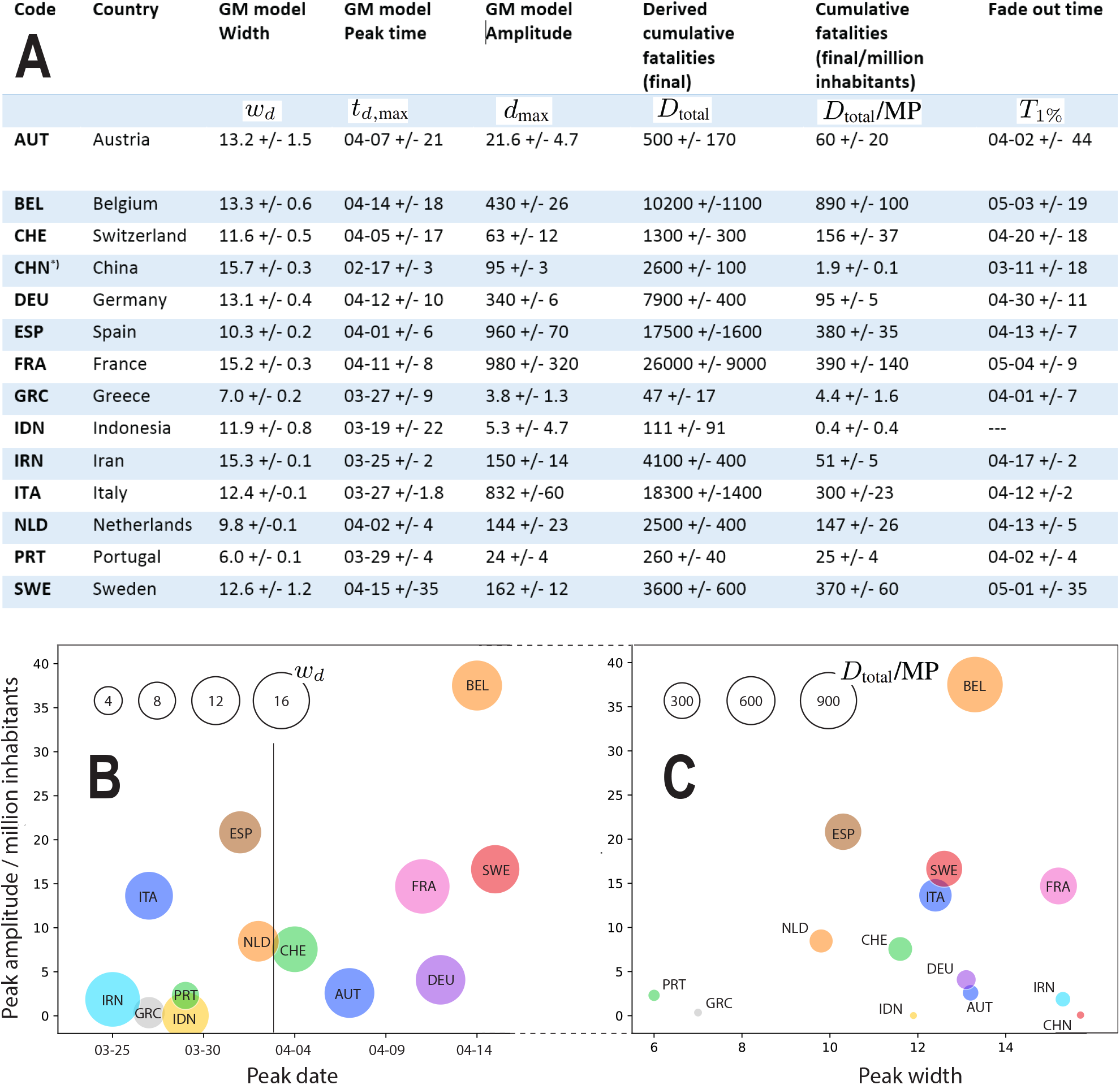
Fitted GM parameters *w*_*d*_, *t*_*d*,max_, and *d*_max_ for those countries, for which sufficient data about fatalities is available by the time or writing. Error bars reported for 95% confidence intervals. **(a)** Table listing the three fitted GM parameters, followed by estimated cumulative number of fatalities *D*total, the same quantity per million people (MP), and the projected dates *Tη* (A10) by which the number of daily *infected* people had reduced to the level of *η* of its maximum value. Number of inhabitants according to OECD [1]. **(b)** Plot of the fitted parameters. The horizontal lines marks the day of this study, April 2nd. China is missing on this plot as its *t*_*d*,max_ occurred on about Feb 17 according to our calculation. **(c)** Peak daily number of fatalities per 1 million inhabitants, i.e., *d*max divided by the number of inhabitants times 10^6. ∗)^ For China we considered only the data during the first pandemic wave, i.e., until a minimum in daily fatalities was clearly reached on March 12.

### D. Additional predictions

Using the GM, one can obtain predictions for the further course of the Covid-19 pandemic analytically from the three descriptors. We here present two possible applications: cumulative fatalities as a function of time and the maximum required number of respiratory equipment as well as its time point.

First, the time evolution of the number of *cumulative* fatalities *D*, plotted in Fig. 5, can be obtained by summing *daily* number of deaths *d*, predicted by our model. In this figure, we rescaled all curves back to normal times so that the future course of cumulative deaths can be easily read-off. One can compute analytically that the date after which the new deaths per day have decreased to 1% of their maximum lies roughly 2 widths after the maximum of daily deaths, and the values, denoted by *T*_*η*_, for each country can be found in Fig. 4(a). Already from visual inspection Fig. 5 suggests for Italy and Spain to plateau first, while France will have to face increasing number of fatalities considerably longer. It also anticipates the cumulative number of fatalities per million people over the entire course of the Covid-19 disease to be highest for France, Spain and Italy.

**Fig 5:**
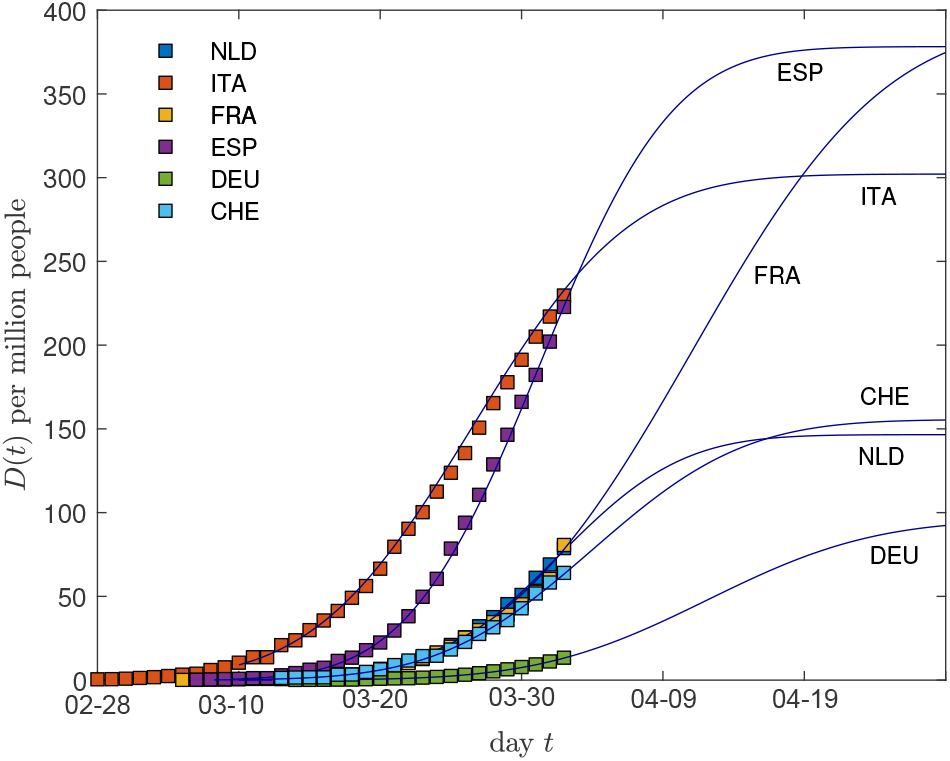
Measured cumulative number of deaths per one million inhabitants (symbols), compared with the GM predictions (A7) for selected fits of Fig. 4. The cumulative number of fatalities determining the height of the future plateau is given by the product of width *w*_*d*_ and *d*max, times 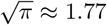.

Next, we estimate the number of required respiratory machines per date for the Covid-19 epidemic. We start by assuming the number of respiratory machines per day to be equal to the cumulative number of *active* seriously sick persons on the given date, where active means not yet recovered by that date. Each new seriously sick person per day (SSP) requires a respiratory machine for some days or even weeks before passing away or recovering from Covid-19. According to other works [19], people to have died from Covid-19 occupied respiratory equipment for an average of 7 days prior to their death, but respiratory equipment may be in use for up to about one month for cases that later recovered. Thus, we may roughly estimate the number of active SSPs per date as the sum of people that became seriously sick within the past 10 days. Please note, however, that we try to only conceptually link the GM to useful quantities, we leave a thorough search of exact numbers to the reader.

As a final step, we need relate SSPs and deaths. Assume that each SSP dies with a constant probability *γ* after some days, i.e. *γ* times SSPS gives the daily fatalities some days ahead. Taking again numbers from Ref. [19], we could use that each deceased patient had used a respiratory machine for an average of 7 days prior to death and thus estimate the daily number of SSPs at a given date by the number of daily fatalities 7 days in the future divided by probability *γ*.

The result of the above estimate reveals that the required number of respiratory equipment itself is a Gaussian curve, roughly centered around the same date as the daily fatality curve, and its peak value is proportional to a multiplicative factor that depends on the width of the Gaussian and ranges between 0.5 and 0.9 for the widths we found, the total number of fatalities *D*_total_, and the ratio of passing away as a SSP *γ*.

## III. Discussion

The preceding section demonstrated how a GM can be used to obtain statistical predictions for a pandemic such as the corona pandemic. The presentation was intentionally conceptual, to convey only the principle idea of such a model. It is clear that such predictions might work better for some countries than others. This is also why the reader should not be distracted too much by the large error in the fitted or derived parameters of Fig. 4.

The question remains, though, how the GM could be justified? In the following we aim to present a number of arguments in favor of the GM that make use of sigmoidal nature of saturating processes such as pandemics and include a numerical argument on the stability of models.

First, let us list some other ‘microscopic’ models that can lead to a Gaussian dynamics of fatalities or infections. One of more prominent ones recently appeared in the Washington Post [14]. Stevens investigated what happens when simulitis spreads in a town, if everyone in the town starts at a random position, moving at a random angle, infecting others upon collision, and recovering after a certain time. The simulated number of infected people rises rapidly as the disease spreads and tapers off as people recover – a bell-shaped curve. We recreated the simulations and found evidence for the applicability of the GM under many circumstances. These results are not reported here, but support our central assumption. From another recent work using a holistic agent-based model [3], where the agents adapt their behavior through artificial intelligence as part of the solution, there seems also evidence from the numerical results presented, that the number of newly infected may be well captured by a Gaussian function.

The key of these behavior lies in the sigmoidal nature of saturating processes. Intuitively, we know that the cumulative number of cases for a wave of any pandemic must start from a constant (often 0), then increase exponentially and eventually saturate at a higher constant level. Functions that capture such a behavior, i.e. a smooth change from a lower constant to a higher constant over a finite duration, are called sigmoidal. The derivative of sigmoidal functions have a bell-shaped form, similar to a Gaussian function, but may be asymmetric in general. We here model the daily fatalities *d*, formally the derivative of the cumulative fatalities *D*. Since we expect cumulative cases *D* to be sigmoidal, from common-sense reasoning as argued above, this fixes the derivative, the daily fatalities *d*, to a bell-shaped form.

Even though all pandemics thus give rise to bell-shaped *d* by this argument, the curve’s parameters might differ, influenced mostly by policy, health system and culture. The predictive power of our model rests on the assumption that these influences are encoded already into the early data of casualties, combined with the assumption that the principal shape of all pandemics is fixed. This is of course an unjustified assumption or, if at all, justified only in a statistical sense when considering large number of trials.

Why do we choose a symmetric bell-shaped form, the Gaussian function? We recognize that other models, such as Poisson functions that fade out slower after their maximum, might be more realistic. However, a symmetric function is the simplest model among all bell-shaped functions and works well enough to convey the idea of such models. Second, the times of greatest interest to policy makers are until the bell-curve’s peak since once passed the health system should be able to cope.

Sigmoidal functions are popular to model saturation processes such as cellular growth [20, 21] or enzyme (Michaelis-Menten) kinetics. Numerous sigmoidal functions exist and they have been used in many variations to predict infectious diseases [8] and have even been linked directly to models such as the SIR model [17]. Not surprisingly, sigmoidal models are frequently used in times of coronavirus [7, 15, 18], most often using a (generalized) logistic function (Richard’s curve). One model particularly similar to our GM [12] also uses the Gaussian integral as sigmoidal function with similar parameters. However, the study only applies to the US, for which we noticed a bimodal Gaussian nature of *d* and thus decided not to model it here. Moreover, the precise methodology remains elusive as of today when we submitted this manuscript, but we assume the author fitted the sigmoidal Gaussian integral function, not its Gaussian derivative. We believe this to be a crucial difference to our procedure, to be discussed in the next paragraph.

A major issue with sigmoidal models is that they are often prone to overfitting [6] and also we in our preliminary experiments found such sigmoidal fits to be sensitive to initial conditions and to often require a large number of parameters. Previously, people have tried to experiment with regularizations [17] to account for such instabilities. Instead, we here choose to fit the logarithm of daily change of cases *d*, not the cumulative cases *D*. The logarithm of *d* weights more evenly values close to the functions maximum and disregards other values. We believe this leads to a more stable model of *d* around its maximum, the turning point of *D*, and the time of interest since most relevant predictions such as peak of the pandemic, time point and width of peak are focused around this maximum.

The above arguments explained our believe in sigmoidal models, but we also see mechanistic problems in other type of models, such as exponentials. Many exponential models rely on doubling times [13], which require intense preprocessing of data, such as smoothing, and are model dependent. Please refer to the methods in the appendix for further discussion of doubling times in the context of the here presented GM. Other exponential models report considerable deviations from an exponential nature, e.g. a power law behavior as the curve flattens [16]. Sigmoidal functions automatically account for exponential growth and subsequent flattening and we thus believe them to be a better predictor.

We must note that we aim to be compared with descriptive models, i.e. models that work only in the statistical limit of large data. In contrast, mechanistic models for infectious diseases [4] are able to study the effect of parameters such as policies, health system or culture on the outcome of a pandemic, and thus to provide more detailed predictions and recommendations.

## IV. Conclusion

The here presented GM allows for simple predictions of future course of the Covid-19 disease and we have provided first evidence that a GM is able to capture the time evolution of the daily fatalities and infections per country. Fitted models describe past data well, including data from China.

The model is so simple that it can be reproduced and applied without detailed knowledge of epidemiology, statistics or programming languages. There are many countries not yet drastically affected by Covid-19, which will likely change for many in the coming weeks, and the GM could for example be used to apply it to such countries as soon as sufficient data is available. Using the recipe presented here, interested readers are in the position to obtain estimates for the shape of the Gaussian curve for their country, state, community, and use this model to compute more quantities of interest, such as our sketch of how to estimate the maximum number of required respiratory machines and the date of this maximum demand. Knowing the time of maximum rush days of SSPs, the maximum number of SSPs and width could help the government and medical agencies in these countries to optimize the managing of the disease wave by appropriate drastic actions for limited time. Moreover, fortunately, as our study here demonstrates, the time of peak of the disease wave differs among countries. Knowledge of these peak times and their durations allows other countries to help those who undergo the peak of the wave at a significantly later time, with breathing apparati and trained medical personal for a brief predictable time.

Besides that we hope to make the public aware of the Gaussian or sigmoidal nature emerging from Covid-19 infections, similar to the numerous discussions of exponential functions in recent times. No pandemic is ever exponential, in the long run it is sigmoidal, and thus makes for a good discussion.

On one hand we are afraid our predictions will become reality, on the other they are more optimistic than all (few) predictions we came across so far. Confronting these predictions and the method with reality will help to either establish or rule out the presented approach within a very short time. It is the simplicity of the model and its missing freedom which will allows us to quickly decide on its usefulness for future applications.

We conclude with a word of caution. We are certainly no experts in this field and a GM is simply a description of a smooth time evolution of deaths. We leave it to the reader to treat the here presented observations and claims with enough care.

Acknowledgements

M.K. acknowledges major contributions by Clarisse Luap (C.L.) to the research of this work. Yéléna F. C. Luap helped creating Fig. 4A. Funding: This work was performed without funding. Author contributions: R.S., F.S. and C.L. motivated, M.K. and R.S. designed the study. M.K. developed the model, evaluated data, created figures. M.K. and R.S. prepared a draft. J.S. carefully restructured and extended the manuscript. All authors contributed to the final manuscript. There are no competing interests to declare. All data is available in either the manuscript or the quoted data repositories.

### A. Methods

In this section we make the concepts used intuitively in the main text rigorous by introducing the necessary mathematical language.

#### Gauss model

Denote the number of daily fatalities as a function of time by *d*(*t*) and the cumulative number of fatalities by *D*(*t*). We model the time evolution of fatalities using the GM, i.e. a Gaussian function of time,

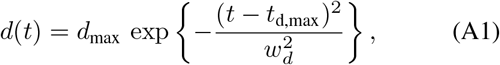

where *w*_*d*_ denotes the width of the Gaussian, *d*_max_ denotes the maximum value of fatalities and *t*_d,max_ the time point at which this maximum is attained. The identical model and notation applies to the number of daily infections *i*(*t*), cumulative infections *I*(*t*) and parameters *i*_max_, *t*_i,max_, *w*_*i*_.

We use publicly available data of *monitored* cumulative death rates *D*_*m*_(*t*), where the subscript *m* is used to distinguish data from our model, to derive the daily death rates by taking the first time derivative

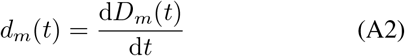

and calculated its natural logarithm ln *d*_*m*_(*t*). The GM dynamics (A1) implies

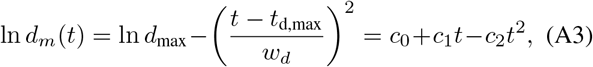

which is a polynomial function of degree 2 with coefficients

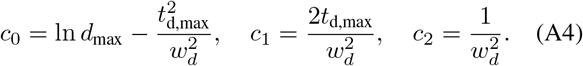

The relevant parameters determining the number of deaths per day, the width of the distribution, as well as the position of the peak are then given by

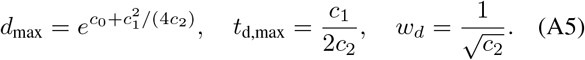

#### Fitting & errors

Using a second order polynomial fit to the data we obtained the coefficients *c*_0_, *c*_1_, *c*_2_ as well as their confidence intervals. For this, the Matlab^R^ function `[P,S,M]=polyfit(t,log(`*I*_*m*_`),2)` on the natural logarithm of the monitored death rates ln *I*_*m*_(*t*) yields the coefficients P=[*c*_2_,*c*_1_,*c*_0_] of the fit as well as information about the confidence intervals. We made use of the function `polyparci` that uses only core Matlab^R^ functions and does not require the Statistics Toolbox. It uses the procedures outlined in the `polyfit` documentation to calculate the co-variance matrix, and uses functions `betainc` and `fzero` to calculate the cumulative t-distribution and the inverse t-distribution for a given probability and degrees-of-freedom. Within the limited amount of time we had to prepare this document, we were unable to compare error estimates from different approaches.

#### Deaths vs. infections

We have applied the same procedure to the measured number of infected people, *i*_*m*_(*t*), giving rise to another set of parameters *i*_max_, *t*_i,max_, and *w*_*i*_. We found that the GM widths for infections *w*_*i*_ and fatalities *w*_*d*_ are similar in magnitude, within errors, and that *τ*_*d*,max_ and *τ*_*i*,max_ differ by a number of days *τ* ≈ 10 [5], that can be considered constant for practical purposes. Our analysis confirms this estimate. It is also useful to introduce the fraction of fatalities among the truly infected (not the reported infected) *f* = *D*_total_*/I*_total_, as this fraction can be expected to vary within limited bounds. We thus write

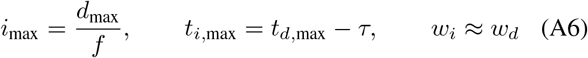

This reduces the number of parameters for a combined study of daily deaths and infections to four, as *f* cannot be considered constant, or further down to three, employing *f* ≈ 5 × 10^−3^ suggested by Fig. 1 of Ref. [5]. We did not make use of these relationships and numbers anywhere in this work, but they can still be used to estimate quantities mentioned below.

While this study mostly focuses on the number of fatalities, we had also included data from 11 countries for the reported number of infections in some of the previous figures that provide evidence for the applicability of the GM. Table I lists the corresponding parameters.

#### Data used

Only countries which as of April 2nd, have reported more than 20 infected or 7 deceased people for more than 10 days. Also, outliers that are better described by a multimodal extension of the GM have been omitted (including the United States) with the exception of China, for which there was a clear end of the first wave on about March 12. This resulted in the 25 counties used here. Using the identical approach, many more countries will be available for analysis within the next few days.

#### Cumulative fatalities

The accumulated number of fatalities at time *t*, which we refer to as cumulative number of fatalities, is the integral of the daily fatalities (A1)

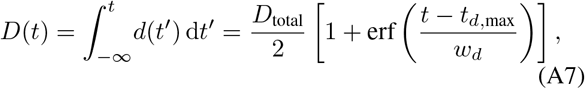

where 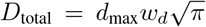 is the projected total number of fatalities at *t* → ∞ and erf is the error function. Using (A7) the time *t*_0_ by which a first patient died from the virus is immediately estimated via *D*(*t*_0_) = 1. Similarly via *I*(*t*_0_) = 1 for the first infected person, so-called patient 0, if one takes into account a time shift *τ* and ratio *f* between *d*_max_ and *i*_max_, cf. (A6), and one ignores the fact that the gaussian is likely to break down in this limit. The explicit expression is *t*_0_ = *t*_*i*,max_ − *w*_*i*_ erf^−1^ (1 − 2*/I*_total_) for the time of first appearance of Covid-19, and this time is specific for each country. Here, erf^−1^ is the inverse error function. For values close to unity it is well approximated by 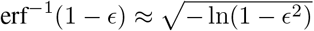.

#### Occupation of respiratory equipment

Most people that died from Covid-19 required respiratory equipment until their death for a period of length *τ*_*r*_ and we assumed this period *τ*_*r*_ to be constant. If *γ* people out of all that require respiratory equipment die, we can estimate the daily occupation of respiratory equipment *r*(*t*) by summing over the past *τ*_*r*_ days of newly seriously sick persons per day (SSPs), which are related to the daily deaths shifted by the typical time *T* from being diagnosed as seriously sick until death. For that, we divide the sum of deaths over the past *τ*_*r*_ days by *γ* to extrapolate to active SSPs at time *t* and hence required respiratory equipment

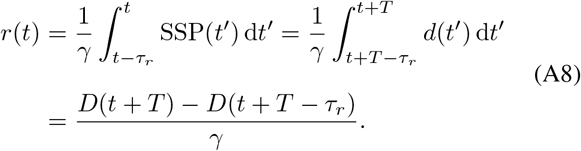

The number of required respiratory machines *r*(*t*) attains its maximum at time *t*_r,max_ = *t*_d,max_ − *T* + *τ*_*r*_*/*2 and thus the peak number of required respiratory machines is *r*_max_ = *r*(*t*_r,max_) = (*D*_total_*/γ*) erf(*τ*_*r*_*/*2*w*_*d*_), where *D*_total_ is the total number of deceased people. This peak *r*_max_ increases with larger occupation times of respiratory machines *τ*_*r*_, larger total number of fatalities *D*_total_ and narrower GM widths *w*_*d*_. For the fitted values of *w*_*d*_ and a *τ*_*r*_ of 10 days, the error function roughly is in the range 0.5 ≤ erf(*τ*_*r*_*/*2*w*_*d*_) ≤ 0.9. Flatten the curve!

#### Percentiles of infection numbers

From *t*_*d*,max_ and *w*_*d*_ we can estimate dates at which time the number of daily infected people *i* will have reduced to the level of *η* ∈ [0, 1] of its maximum value. These times denoted as *T*_*η*_ are given by

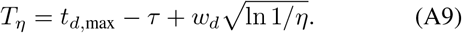

For *η* = 1% and *η* = 1‰ these times are explicitly given, employing the typical delay time *τ* ≈ 10 days (A6), by

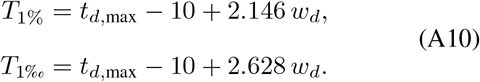

The corresponding dates are listed in Fig. 4. It is also possible to estimate dates, for which less than a certain *η* of the total population remains infected and potentially dangerous to initiate another outbreak. This time is given by

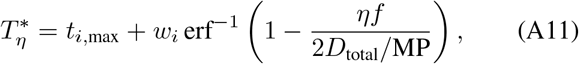

where *D*_total_*/*MP had been tabulated (Fig. 4), erf^−1^ is the inverse error function, and *f* defined by (A6) may be approximated by the value mentioned there. In this expression *t*_*i*,max_ and *w*_*i*_ can also be approximated by using the values calculated from fatalities, as described already. For *η* = 10^−6^ (one per million inhabitants), and a typical 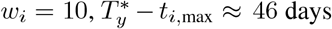 for *D*_total_/MP = 100.

#### Doubling times

Doubling times, here denoted by *k*, are used to characterize the strength of an exponential growth process, independent of the exponential amplitude. A doubling time quantifies the time span required for the exponential to double (or, up to convention, to have doubled). Assuming a purely exponential growth, both *d*(*t*) = *d*_max_ exp(*νt*) and *D*(*t*) = *d*(*t*)*/ν* increase mono-exponentially with time, and the doubling time *k* is a constant, *k* = *ν*^−1^ ln 2, while *ν* = *d*^*l*^(*t*)*/d*(*t*) = *D*^*l*^(*t*)*/D*(*t*). For the GM the doubling time based on *d*(*t*) is thus given by

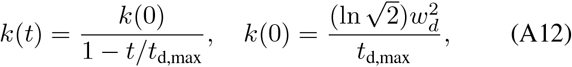

while the doubling time based on *D*(*t*) is given by *k*(*t*) = (ln 2)*/*[*k* ln *D*(*t*)*/dt*] = (ln 2)*D*(*t*)*/d*(*t*). It is thus easy to calculate two versions of doubling times with the GM parameters at hand, using either daily or total measures, which differ if the growth is not ideally exponential. While doubling times are convenient as they alter only weakly during exponential growth, they are difficult to extract from data directly without applying smoothing procedures that differ from publication to publication, and they are not uniquely defined. For this reason we do not recommend to proceed with an analysis on reported doubling times, as done in [13], unless the raw data is unavailable.

## Data Availability

All data used in this study is available online to the public.

https://pomber.github.io/covid19/timeseries.json

https://data.oecd.org

---

**TABLE I:** GM parameters *w_i_*, *t*_*i*,max_, and *i*_max_ for those countries, for which sufficient data about infections, but insufficient data about fatalities is available to us, as of April 2. The coefficient *i*_max_ we could not extract from the existing data with an error less than 100%.^‥^Parameters *w*_*i*_, *t*_*i*,max_, *i*_max_ were used to create Fig. 3.

| $\alpha_3$ code | Country | $w_i$ | $t_{i,\max}$ | $i_{\max}$ | $I_{\text{total}}$ | $I_{\text{total}}/\text{MP}$ |
| --- | --- | --- | --- | --- | --- | --- |
| BRA | Brazil | $11.1 \pm 0.2$ | $04-03 \pm 8$ | $800 \pm 350$ | $16000 \pm 7000$ | $78 \pm 35$ |
| CHL | Chile | $11.7 \pm 0.2$ | $04-02 \pm 7$ | $321 \pm 33$ | $6600 \pm 800$ | $370 \pm 45$ |
| GBR | Great Britain | $20.0 \pm 0.7$ | $04-27 \pm 14$ | — | — | — |
| JPN | Japan | $38.0 \pm 2.3$ | $04-27 \pm 22$ | $195 \pm 90$ | $13000 \pm 7000$ | $104 \pm 54$ |
| SAU | Saudi Arabia | $15.4 \pm 0.6$ | $04-02 \pm 13$ | $140 \pm 23$ | $3900 \pm 700$ | $120 \pm 24$ |
| SRB | Serbia | $10.3 \pm 0.3$ | $04-01 \pm 8$ | $113 \pm 50$ | $2100 \pm 1000$ | $290 \pm 130$ |
| PAK | Pakistan | $10.3 \pm 0.3$ | $03-28 \pm 11$ | $170 \pm 80$ | $3200 \pm 1600$ | $16 \pm 8$ |
| PER | Peru | $16.8 \pm 1.3$ | $04-09 \pm 28$ | $148 \pm 64$ | $4400 \pm 2300$ | $140 \pm 70$ |
| POL | Poland | $15.0 \pm 0.4$ | $04-07 \pm 10$ | $340 \pm 50$ | $9100 \pm 1500$ | $240 \pm 40$ |
| ROU | Romania | $18.9 \pm 1.3$ | $04-19 \pm 26$ | $690 \pm 90$ | $23200 \pm 4600$ | $1200 \pm 250$ |
| USA | United States | $14.8 \pm 0.2$ | $04-14 \pm 5$ | — | — | — |

